# Patient associations as knowledge intermediaries in breast cancer governance in Mali: a prospective qualitative analysis

**DOI:** 10.64898/2026.05.11.26352883

**Authors:** Julie Robin, Madani Ly, Hamidou Niangaly, Clémence Schantz, Abdelaye Keita, Kadiatou Kanté, Valéry Ridde, the Senovie group

**Affiliations:** Centre Population et Développement (UMR 196), Université Paris Cité, Institut de Recherche pour le Développement, Université Sorbonne Paris Nord, Inserm, F-75006 Paris, France; Forum Médical, Centre International d’Oncologie (CIO), Bamako, Mali; Département Études et recherches, Institut National de Santé Publique, Bamako, Mali; Laboratoire d’Epidémiologie des Maladies Chroniques et Neurologiques (LEMACEN), Ecole Doctorale des Sciences de la Santé, Université d’Abomey Calavi, Cotonou, Benin; Institut National de Santé Publique, Bamako, Mali; Association « Les Combattantes du Cancer », Bamako, Mali

## Abstract

**Background:** Breast cancer is an increasing public health concern in many low- and middle-income countries, yet prevention and care decisions do not consistently rely on evidence. In Mali, patient associations are increasingly visible in cancer advocacy, but their potential role in mediating research and experiential knowledge within decision-making remains poorly understood. This study adopts a systemic perspective on knowledge transfer to examine the conditions under which a patient association–led knowledge intermediation mechanism could plausibly emerge and be embedded within breast cancer governance.

**Methods:** We conducted a qualitative pre-implementation study based on 29 semi-structured interviews, one focus group, and participant observation involving patient association members and leaders, health professionals, researchers, decision-makers, international partners, and media representatives. Data were analysed using an adapted version of the Consolidated Framework for Implementation Research to guide construct-based coding, followed by cross-domain analysis to identify clusters of interacting determinants synthesised into broader analytical configurations.

**Results:** Analysis identified three configurations shaping the plausibility of patient association–led knowledge intermediation in breast cancer governance, across institutional, organisational, and epistemic dimensions. First, fragmented institutional arrangements, project-based financing, and discontinuous coordination limit the stabilisation of interfaces and continuity of interactions, thereby constraining the emergence of sustained knowledge transfer processes.

Second, patient associations hold strong experiential legitimacy and mobilisation capacity, but their ability to assume intermediation roles is constrained by stigma, limited confidence in engaging with scientific and policy-relevant knowledge, restricted access to data and analytical resources, and weak integration into institutional decision-making spaces. These constraints are also shaped by gendered illness trajectories and social expectations that affect women’s ability to engage publicly and the recognition of experiential knowledge as policy-relevant evidence. Third, competing framings of cancer problems, combined with institutional norms privileging biomedical and quantitative evidence, narrow the range of knowledge considered actionable, limiting the integration of experiential and contextual knowledge into decision-making. Together, these configurations identify the interdependent conditions shaping the emergence, functioning, and durability of association-led knowledge intermediation mechanism. Without simultaneous alignment of institutional, organisational, and epistemic conditions, intermediation is likely to remain partial, episodic, and weakly sustained.

**Conclusion:** These findings suggest that association-led KT cannot rely on patient associations alone, nor on capacity-building alone. It requires implementation strategies that build structured interfaces enabling patient associations, researchers, clinicians, and decision-makers to jointly interpret and translate heterogeneous forms of knowledge into actionable inputs, while addressing the gendered conditions of women’s engagement and the uneven recognition of experiential knowledge in institutional evidence use. Further research is needed to test how such relational configurations can be operationalised and sustained in constrained health systems.

## Introduction

Breast cancer is an increasing public health concern in low- and middle-income countries (LMICs), where late diagnosis, limited access to treatment, and fragile health system responses contribute to high mortality and substantial social and economic consequences (1). Despite this growing burden, cancer has long remained weakly prioritised in many LMIC health agendas and funding allocations, which have historically focused on maternal and child health and infectious diseases (2,3). This misalignment has been associated with weak institutional anchoring of cancer control, unstable financing, and limited strategic coordination (2,3).

This situation also reflects broader difficulties in how evidence is produced, circulated, and used in health decision-making. Over the past two decades, international commitments have repeatedly called for more evidence-informed approaches to cancer control and for stronger links between research, data systems, and policy-making (4–6). However, empirical analyses increasingly show that these commitments have translated unevenly into practice. Two decades after the World Health Organization’s first calls for evidence-informed cancer control, analyses of national cancer control plans in LMICs point to incomplete integration of cancer-related data, weak articulation between evidence, priority-setting and implementation, and limited institutionalisation of evidence use beyond planning documents (7).

In parallel, international and national frameworks increasingly recognise the value of lived experience and the role of patients and civil society in health decision-making, including in cancer control (8,9). Yet, patients’ participation remains weakly institutionalised and often symbolic, with limited influence over priority-setting or resource allocation (10). In cancer control, patient associations are frequently mobilised for awareness-raising and advocacy, but rarely occupy sustained roles in evidence use or strategic decision-making (7,11). This raises the question of under what conditions they might play a more substantive role in the circulation and use of knowledge.

Knowledge transfer (KT) research provides a useful lens for addressing this question. It shows that evidence use is highly context-dependent and depends not only on the availability of knowledge, but also on the actors, relationships, and organisational and political conditions that shape its circulation and uptake (12–17). This literature also highlights the importance of intermediary functions in supporting exchanges across research, practice, and policy arenas (18–20). In this perspective, intermediation is understood not as a predefined actor role, but as a set of functions and relational practices—such as translation, sense-making, and relationship-building—that may be carried by different individual or collective actors depending on the context (21). This question is particularly salient in cancer control, where decision-making requires combining biomedical, epidemiological, economic, and experiential forms of knowledge (22–25).

In this context, patient associations occupy a strategic position at the intersection of lived experience, care practices, and public debate. However, the conditions under which they can act as credible and influential knowledge intermediaries remain insufficiently documented, particularly in LMICs. This study adopts a pre-implementation perspective to examine the conditions under which an association-led knowledge transfer mechanism around breast cancer could plausibly emerge, function, and consolidate in Mali.

## Methods

### Study design

We conducted a qualitative study using semi-structured interviews to examine the conditions under which an association-led KT mechanism on breast cancer could plausibly emerge in Mali. The mechanism under consideration is prospective and positions patient associations as central intermediary actors within a broader intermediation arrangement involving researchers, health professionals, and decision-makers.

### Conceptual framework

This study was informed by an adapted version of the Consolidated Framework for Implementation Research (CFIR 2.0)(26) to examine factors shaping the anticipated implementation of a patient association–led KT mechanism in the malian cancer context. CFIR is a determinant framework designed to identify multilevel factors influencing implementation processes, as distinguished from process models or grand theories in implementation science (27). It was selected for its capacity to structure multi-level determinants across innovation characteristics, inner and outer settings, individual actors, and implementation processes. The emphasis on contextual determinants aligns with implementation science scholarship highlighting the centrality and multidimensional nature of context in shaping implementation outcomes (28). Given the central role of women’s lived experience in breast cancer patient associations, the analysis remained attentive to gender as a cross-cutting dimension (29,30).

The CFIR framework was prospectively adapted through a structured, theory-informed process drawing on literature on KT in health, civil society engagement, and cancer governance in low- and middle-income countries. Construct selection and refinement were guided by relevance to knowledge mediation and to the hypothesised intermediary role of patient associations. Full details of the adaptation process are provided in S1 File.

Constructs were selected, merged, or re-labelled to improve contextual fit and analytical clarity. The adapted framework was used to guide interview development and first-cycle coding, while allowing inductive refinement during analysis. Detailed procedures for construct adaptation and coding are provided in S1 File.

### Operationalisation of CFIR domains

CFIR domains were operationalised prospectively to examine anticipated implementation conditions of a patient association–led KT mechanism in Mali. The innovation was conceptualised as a hypothesised intermediary arrangement through which patient associations could mediate research-based and collectivised experiential evidence into decision-making arenas; the SENOVIE project, which generated part of the research evidence used in this study, was treated as an external support through its role in evidence production and provision of limited resources for KT. The inner setting focused on patient associations’ organisational capacity and routines; the outer setting captured Mali’s cancer governance environment and evidence-use context. Individual-level determinants were analysed across patient representatives, clinicians, researchers and decision-makers (e.g., perceived legitimacy and confidence with evidence), and process constructs were used to examine anticipated enactment and coordination dynamics (e.g., engagement and feedback). Further details are provided in S1 File.

In this study, patient experiential knowledge refers to the insights and understandings developed through living with illness, treatment, and interactions with healthcare systems (31).

### Data collection

Semi-structured interviews were conducted between 16 April 2024 and 8 July 2025. Data collection spanned a period marked by institutional continuity in cancer governance; no major structural reform occurred during this time. Interviews explicitly explored both past and current dynamics to account for temporal variation.

In total, we conducted 29 individual interviews with actors involved, in different capacities, in the production, mediation, or use of knowledge related to breast cancer in Mali. Participants included 14 representatives of patient associations and civil society organisations (including NGOs acting as technical partners to patient associations), 6 health policy decision-makers, 4 representatives of research institutions, 4 health professionals and researchers involved in breast cancer care or research, 1 representative of an international organisation, and 1 journalist with recognised experience in reporting on health-related issues.

Participants were selected using purposive sampling, a strategy commonly used in qualitative and implementation research to identify information-rich cases based on their roles and positions within the system (32). Sampling aimed to ensure representation of actors occupying distinct positions in the production, mediation, and use of breast cancer-related knowledge. Initial participants were identified through institutional mapping and SENOVIE networks, with subsequent recruitment guided by emerging analytical needs.

All interviews were conducted in person and in French. Recorded interview duration ranged from 17 to 92 minutes, with a median duration of 56 minutes. Most interviews lasted around one hour. Interviews were audio-recorded with participants’ verbal informed consent when permitted and transcribed verbatim for analysis. Detailed notes were taken for non-recorded interviews.

Recruitment continued until sufficient diversity of perspectives was achieved and no substantively new analytical dimensions emerged across interviews.

In addition to individual interviews, one focus group discussion was conducted with 17 members of *Palissade*, a platform bringing together patient associations working on non-communicable diseases (NCDs) in Mali . The focus group aimed to explore collective perceptions regarding the role of patient associations in knowledge mediation and interactions with public institutions, and to contrast cancer-related dynamics with those observed in other NCDs. Participants were selected based on their active involvement in association leadership and platform coordination. The focus group was audio-recorded with participants’ verbal consent and transcribed verbatim.

In addition to interviews, the study drew on sustained participant observation conducted within two complementary settings. First, participant observation was carried out as part of the SENOVIE project activities between October 2022 and October 2025, including steering committee meetings, scientific committee meetings, conferences, and participatory workshops. Second, participant observation was conducted between October 2024 and October 2025 within the activities of the breast cancer patient association Les Combattantes du Cancer, including awareness-raising activities, public conferences, and communication initiatives on social media.

These observations were embedded in ongoing project and association activities. They were documented through field notes and used to contextualise interview data, inform the interpretation of organisational, relational, and legitimacy dynamics, and support analytical triangulation. Participant observation was not treated as a standalone data source, but as complementary material contributing to analytical depth and reflexivity.

### Data analysis

Data were analysed using qualitative content analysis combining deductive and inductive approaches. An adapted version of the CFIR framework guided first-cycle coding of the full corpus. Transcripts were managed and coded using NVivo (QSR International).

Following initial coding, constructs were reviewed and refined to ensure coherence with the study’s pre-implementation perspective. Inductive codes were allowed to emerge beyond pre-selected constructs and new determinants were retained when they appeared recurrent across interviews.

The analysis proceeded through three iterative stages. First, findings were synthesised at the level of individual CFIR constructs. For each construct, its relative empirical support was assessed qualitatively, based on recurrence across interviews, diversity of respondent profiles, and convergence of accounts. This step ensured that subsequent interpretations remained grounded in construct-level evidence. Second, a cross-construct analysis was conducted to identify recurrent patterns of co-occurring determinants across interviews. Rather than treating constructs as independent factors, we examined how they interacted within participants’ narratives. These patterns were stabilised into empirical clusters of interacting determinants, capturing recurring combinations of conditions shaping knowledge use and intermediation.

Third, these clusters were interpreted at a more integrative level to identify broader configurations. We use the notion of configuration as an analytical device to synthesise recurrent cross-construct patterns in the data and to highlight how multiple determinants appear to combine in shaping the conditions for knowledge intermediation (33). Configurations were therefore not derived from isolated constructs, but from recurring patterns of interaction between constructs across participants’ accounts. This analytical step did not follow a formal configurational method such as qualitative comparative analysis or realist evaluation, but used configuration in a modest interpretive sense as an additional level of analysis alongside CFIR constructs.

Focus group data were analysed using the same logic and integrated to refine cross-domain interpretations. Field notes were not coded as an independent dataset but were used to refine construct interpretation and support triangulation. Constructs were not treated as belonging to mutually exclusive categories. Some determinants appeared across several empirical clusters, reflecting the fact that the same structural conditions could contribute to different cross-construct interpretations depending on how they interacted with other factors in actors’ accounts. S2 File therefore indicates where constructs contributed to more than one configuration, illustrating the partially overlapping nature of the arrangements shaping the anticipated feasibility of intermediation.

Detailed coding procedures, construct refinement processes, and criteria for assessing empirical support are described in S1 File. A detailed mapping of constructs to configurations is provided in S2 File. To enhance analytical transparency while preserving the readability of the results section, illustrative quotations supporting each configuration and cross-construct pattern are provided in S3 File.

### Researcher reflexivity and methodological rigor

This study is reported in accordance with the Consolidated Criteria for Reporting Qualitative Research (COREQ) checklist. All interviews were conducted by the first author, whose positioning and long-standing experience in health systems research in Mali informed reflexive attention throughout the study; analytical rigor was supported through triangulation, regular discussions within the research team, and an audit trail, with further details provided in S4 File.

## Results

The analysis revealed three interrelated configurations, identified through recurrent cross-construct patterns across interviews: (i) the conditions under which evidence use may become routinised within institutional decision-making processes; (ii) the capacity of patient associations to consolidate a credible intermediary role beyond individual testimony; and (iii) the contours of a legitimate and actionable evidence perimeter able to navigate competing problem framings. These configurations should not be understood as mutually exclusive categories, but as recurrent analytical patterns capturing how interacting determinants combine to shape different dimensions of the plausibility of intermediation.

### Configuration 1: Discontinuous knowledge use in a project- and person-dependent governance environment

This configuration reflects a recurrent pattern in which cancer-related action and knowledge use are shaped by discontinuous institutional attention within a fragmented governance environment. Initiatives emerge through projects, individual leadership, or external pressure, but rarely accumulate over time. As a result, both research-based and experiential knowledge circulate in unstable and non-cumulative ways, limiting their contribution to sustained decision-making processes.

The central issue in this configuration is the stabilisation of knowledge circulation, uptake, and reuse over time. The feasibility of intermediation therefore depends on whether continuity can be created across actors, projects, and policy moments within a governance system that remains weakly institutionalised. Two recurrent cross-construct patterns are particularly salient within this configuration.

#### Pattern 1 : Discontinuous knowledge circulation in a fragmented governance environment

Across interviews, cancer governance is consistently described as fragmented and weakly anchored institutionally. Responsibilities for cancer-related research, planning, and decision-making are dispersed across multiple institutions, without any actor or body effectively coordinating the field. Respondents describe a combination of factors contributing to this discontinuity: successive institutional reforms blurring mandates, weak institutional memory, limited financial resources dedicated to cancer coordination, dependence on project funding, and weak stabilization of responsibilities.

This governance arrangement appears to make attention to cancer largely dependent on individual or opportunistic dynamics. When key individuals move on or projects end, issues are described as quickly falling off institutional agendas. Several interviewees explicitly contrast this situation with infectious disease programmes, which are seen as benefiting from dedicated structures, clearer leadership, and routine follow-up mechanisms.

This fragmentation is described as particularly pronounced in the field of non-communicable diseases. In the cancer field, initiatives are characterised as isolated and highly dependent on individual clinicians, reflecting the absence of clear institutional leadership. This limits continuity and hinders the consolidation of cancer as a sustained policy priority.

Within this fragmented governance landscape, decision-making processes are also described as weakly connected to research production. Respondents highlight the absence of formal routines or recognised interfaces linking researchers, hospitals, and decision-makers.

Research is described as being conducted within institutions, but with limited circulation of findings. Decision-makers themselves acknowledge limited awareness of ongoing research activities, including those produced within their own services.

Patient associations are described as remaining insufficiently involved in decision-making arenas, despite some recent improvements linked to ad hoc initiatives. More broadly, not all clinical specialties involved in cancer care are systematically included in key coordination moments led by the Ministry, limiting both the diversity of contributions and the circulation of knowledge within decision spaces.

Consequently, research evidence is often described as existing “on paper” but rarely mobilised in practice. Reports are said to accumulate without being revisited, synthesised, or translated into decisions.

Evidence use therefore depends on individual initiative or short-lived opportunities, and circulates in short and fragile cycles tied to individuals or projects, with limited possibility of cumulative use over time.

This pattern corresponds to recurrent cross-construct combinations identified across interviews (C1-A to C1-C, S2 File), including institutional fragmentation, absence of formalised interfaces, weak demand for evidence, constrained circulation between research and decision-making spaces and reliance on interpersonal coordination. Combined, they point to predominantly informal and discontinuous pathways of knowledge circulation, preventing the cumulative and institutionalised use of both research-based and experiential knowledge.

#### Pattern 2. Short-lived policy windows and repeated loss of momentum

Cancer control is widely described as chronically underfunded within domestic health budgets. Public investment is perceived as limited and discontinuous, with resources concentrated on a small number of curative components, while prevention, early diagnosis, research coordination, and planning received little sustained support.

In this context, most cancer-related initiatives rely on external, project-based funding. These projects create temporary windows of action but rarely lead to long-term institutionalisation as they remain weakly embedded in routine planning, budgeting, and accountability mechanisms.

The HPV vaccination programme was repeatedly cited as a notable exception, benefiting from strong international mobilisation and integration into existing immunisation structures. Beyond this case, policy attention to cancer is described as largely contingent on external pressure or individual leadership.

Respondents nevertheless point to a growing interest among some decision-makers in locally produced cancer data, which has periodically opened opportunities for dialogue and interaction, notably through participatory workshops linked to the development of a national cancer plan and to broader non-communicable disease strategies.

Interviewees referred to the Coordination and Monitoring Committee for the Fight Against Cancer in Mali as an attempt to structure coordination at central level, but reported that its activities stalled shortly after its establishment due to the absence of dedicated financial resources. More recently, several respondents referred to recent policy discussions initiated under specific ministerial leadership, following mobilisation by patient associations.

Institutional instability, limited financing, and frequent turnover are perceived as contributing to repeated losses of momentum. This was reflected in the limited implementation and follow-up of the national cancer control plan developed in 2008, as well as in the stalled progress of a subsequent draft initiated in 2023 according to interviewees. A similar pattern was also visible in the adoption of Ordinance No. 2013-023/P-RM of 3 December 2013, formally establishing a National Oncology Centre. Although created by decision of the Council of Ministers, the Centre was not effectively operationalised in the years that followed, illustrating the gap between formal institutional recognition and sustained organisational implementation.

Several respondents also pointed to repeated losses of institutional capacity following external or individual investments in cancer-related services. Technical platforms, clinical practices, and specialised expertise were described as having been introduced, supported, or strengthened either through external partnerships or through the initiative of individual health professionals within particular services or hospitals, but often struggling to endure once this support weakened, priorities shifted, or the individuals concerned moved on. Interviewees referred to several such examples, including the development of anatomic pathology services, the introduction or support of breast micro-biopsy techniques, and initiatives aimed at assessing or improving selected practices. These examples were not discussed as isolated cases, but as illustrations of a broader pattern in which gains remain difficult to stabilise in the absence of durable institutional arrangements. Respondents described this dynamic as contributing over time to losses of organisational memory, technical continuity, and cumulative capacity. Nevertheless, trajectories such as Weekend-70 suggest that more sustained and scalable effects can be achieved when committed leadership is combined with substantial external support, including project funding, training, equipment, and communication activities. However, such cases also underline the extent to which continuity itself may remain dependent on external financial and technical backing rather than on fully institutionalised domestic arrangements. Overall, these trajectories suggest that the issue is not external or individual support per se, but the difficulty of transforming such support into cumulatively anchored institutional capacity.

These patterns correspond to recurrent cross-construct combinations identified across interviews (C1-D and C1-E, S2 File), including project-based financing, dependence on individual leadership, weak domestic investment, institutional instability, and limited follow-up mechanisms. Together, these findings point to short-lived windows during which knowledge may circulate temporarily, without becoming durably embedded in planning, budgeting, or monitoring systems. Several respondents described a tendency for actors to re-engage in new initiatives without systematically building on previous experiences, contributing to fragmented and discontinuous trajectories of action and knowledge use across the system.

Across these two patterns, patient associations appear as partial carriers of continuity within a system where institutional coordination and follow-up remain weak. Several respondents highlighted the role of patient associations in public mobilisation, awareness-raising, and selected concrete advances, including the 2009 decree introducing free chemotherapy, often cited as a major milestone to which associative mobilisation is perceived to have contributed, alongside broader political and institutional dynamics. Associations have also facilitated partnerships with international actors, enabling specific improvements in equipment or services. However, these contributions remain largely dependent on external funding, favourable political opportunities, or individual initiatives. They are rarely embedded in long-term planning, budgeting, and monitoring mechanisms.

In this configuration, the recognition of associations appears less as a sign of stable institutional integration than as part of a compensatory dynamic within a system where coordination and routine knowledge use remain weak. Associations help sustain action where institutions struggle to anchor it over time, while without substituting for more structured arrangements.

Discontinuity is linked not only to fragmented governance structures but also to the temporary and person-dependent character of policy attention itself. In this configuration, the stabilisation of intermediation remains constrained by the limited continuity, interfaces, and follow-up arrangements through which knowledge could be cumulatively reused. Knowledge and experience generated through successive initiatives therefore appear to be repeatedly reset rather than consolidated, limiting their institutionalisation within governance processes.

### Configuration 2: Conditional intermediation shaped by recognised experiential legitimacy and limited organisational consolidation

In this configuration, the central issue is not legitimacy per se, but the extent to which this legitimacy can be translated into stabilised organisational capacities for intermediation. The feasibility of intermediation therefore depends on the consolidation of collective routines, access to data, and arrangements for aggregating and translating experiential knowledge. Two recurrent cross-construct patterns are particularly salient within this configuration.

#### Pattern 3. Experiential legitimacy and visible mobilisation without stabilised knowledge aggregation

Interviews data suggest that patient associations are widely recognised for their mobilisation and advocacy role, with repeated references to concrete achievements and strong symbolic visibility. Building on this recognised role, they are described as central actors in sustaining the visibility of cancer in the public space, particularly through recurring events such as Pink October and World Cancer Day. They are actively involved in awareness-raising, public testimony, psychosocial support, and advocacy around access to care and treatment costs, positioning themselves as complementary actors focused on making cancer visible and giving voice to patients. This recognition is further reinforced by examples from other disease areas, which contribute to consolidating the broader legitimacy of patient associations within the health sector.

Futhermore, their participation is formally recognised in policy discourse and in the Malian Hospital Law framework, notably through the Patient Charter (34), which codifies fundamental patient rights within health facilities. However, this recognition remains largely formal, as these provisions are only weakly known and rarely mobilised in practice by both healthcare professionals and patient associations.

However, this visibility and formal recognition do not in themselves translate into a sustained capacity to engage in knowledge intermediation. This legitimacy is largely grounded in lived experience of illness, rooted in trajectories of the body, care, and social relations, and carries strong moral authority and social resonance. Yet it appears unevenly recognised across domains of action, being more readily valued in awareness-raising and advocacy than in processes of knowledge production, interpretation, and policy-oriented engagement.

As a result, the knowledge produced and circulated by associations remains predominantly experiential and weakly stabilised. Advocacy narratives are largely framed around social suffering, access barriers, and the financial burden of cancer care, generating situated insights into care trajectories, treatment discontinuities, and the practical frictions patients encounter when navigating the health system. What remains limited is not the existence of such knowledge, but its aggregation, formalisation, and translation into institutionally usable forms.

Testimonial knowledge remains largely individual and difficult to consolidate, while weak communication and coordination within and between associations further constrain collective learning. These limitations are reinforced by organisational fragmentation and dependence on individual actors, preventing the stabilisation of intermediary functions over time.

This pattern corresponds to recurrent cross-construct combinations identified across interviews (C2-A and C2-B, S2 File), including experiential legitimacy, social recognition, mobilisation capacity, organisational fragmentation, and dependence on individual actors. These elements converge to suggest that experiential knowledge circulates through mobilisation and testimony while remaining weakly aggregated and institutionalised, limiting its translation into collectively validated and decision-oriented forms. Overall, these findings suggest that the engagement of patient associations is structured by a form of experiential legitimacy that is both enabling and constraining, constituting a central element of Configuration 2.

#### Pattern 4. Constrained access to data and limited capacity to engage with evidence and institutions

Associations recognise the need for evidence and structured knowledge to influence decision-making but face major constraints in accessing data, tools, and institutional codes. Several respondents explicitly note that participation in strategic or policy spaces requires data, evidence, and familiarity with institutional norms. However, association members consistently express low confidence in their ability to engage with scientific or policy-relevant knowledge. Limited access to data, statistics, and analytical tools emerged as a central constraint. Associations describe being invited to speak in public or institutional spaces without having relevant data readily available, which undermines both their confidence and credibility. Under these conditions, research-based evidence is perceived as necessary but remains largely external to associations’ practices, mobilised only sporadically through individual leaders or external partners.

This gap produces a form of restraint: associations are aware of what would be required to engage credibly with evidence, but do not feel equipped to do so. Rather than indicating an intermediary role exercised through other means, this reinforces the perception—shared by associations themselves—that such a role remains difficult to sustain under current conditions.

These constraints are further reinforced by how associations are positioned within decision-making and professional arenas. Respondents describe them as frequently invited by authorities for visibility, mobilisation, or symbolic representation, while remaining largely excluded from technical or strategic discussions. Even when formally present, participation is often described as superficial, with little expectation that associations would contribute substantively to deliberations. More broadly, limited familiarity with institutional processes and governance mechanisms constrains their ability to translate experiential knowledge into structured and actionable proposals. In practice, associations tend to be visible during symbolic moments but remain marginal in sustained policy processes.

Structural organisational fragility further limits their capacity to engage in intermediation. Financial resources are scarce and unstable, participation depends on voluntary engagement, and health-related constraints affect continuity and leadership renewal. Several respondents describe associations as operating with very few active members due to treatment, recurrences, and advanced disease. This fragility is not solely organisational but is shaped by the gendered conditions under which women experience illness and engage in collective action, including care responsibilities, social expectations, physical and emotional burden of disease trajectories and the broader social consequences of disease trajectories for women’s family roles, marital relationships, community belonging, and public visibility (30).

In addition, access to sustained guidance, mentoring, and training adapted to the needs of patient associations remains limited. Training opportunities are described as sporadic and unevenly distributed, while language barriers further restrict access to regional and international knowledge resources.

Stigma associated with cancer is frequently reported as a barrier to engagement, particularly for women, contributing to reluctance, withdrawal, or selective participation. However, these dynamics are not uniform. Some women describe processes of reappropriation of their bodies and illness experience, including forms of bodily exposure, humour, and collective support, through which stigma could be partially resisted or transformed. These practices reflect forms of individual and collective reconstruction, without necessarily challenging underlying gender norms. These findings highlight an important tension: while stigma constrains engagement, it may also, in certain cases, generate situated forms of re-engagement and visibility.

These elements corresponds to recurrent cross-construct combinations identified across interviews (C2-C and C2-D, S2 File), including limited access to data, low analytical capacity, weak understanding of institutional processes, organisational fragility, and constrained participation conditions. These cross-construct patterns suggest that the translation of experiential knowledge into credible, structured, and institutionally usable inputs remains difficult under current conditions. They also point to a persistent gap between institutional expectations for evidence-based contributions and the forms of knowledge that associations are currently able to produce and mobilise. Across these two patterns, patient associations occupy an ambiguous position within knowledge processes. Their experiential legitimacy supports visibility and mobilisation, while their involvement in sustained decision-oriented knowledge processes remains constrained by limited organisational consolidation. In this configuration, intermediation appears conditional, situational, and dependent on individual actors and external support more than on institutionalised organisational capacities. This limits the continuity and cumulative effects of their contributions to knowledge circulation.

### Configuration 3: Negotiated boundaries of evidence and the conditions of intermediation

In this configuration, the central issue is not only the availability of evidence, but the criteria through which evidence is recognised, prioritised, and used in decision-making. The feasibility of intermediation depends on the capacity to align, translate, and negotiate across competing evidentiary expectations. Two recurrent cross-construct patterns are particularly salient within this configuration.

#### Pattern 5. Competing problem framings and divergent expectations toward evidence

Respondents tend to frame cancer through different lenses: decision-makers most often emphasise budgetary constraints, health professionals clinical issues, and patient associations social and moral dimensions.

Decision-makers primarily approach cancer through a budgetary and cost-containment lens, focusing on treatment affordability and the State’s capacity to sustain financial commitments. Healthcare professionals frame cancer mainly as a complex clinical condition requiring specialised expertise, equipment, and technical decisions, while patient associations frame it as a social and moral issue, emphasising catastrophic expenditures, access barriers, suffering, and injustice.

These framings coexist without a shared evidentiary reference, contributing to parallel debates and unstable prioritisation. Beyond these competing framings, respondents highlight structural features that further complicate cancer’s positioning within decision arenas.

The combination of high technical complexity and limited professional critical mass contributes to cancer’s fragile positioning in multi-sectoral decision arenas. In contexts where multiple health priorities compete, the numerical and institutional weight of actors influences which problems and which forms of evidence gain traction.

The absence of formalised interfaces between actors and the weak articulation between experiential and scientific knowledge further limit the emergence of a shared evidentiary reference. Research-based evidence and experiential knowledge are not contested in principle, but are assessed through distinct and often incompatible lenses.

This pattern corresponds to recurrent cross-construct combinations identified across interviews (C3-A and C3-B, S2 File), including divergent framings, absence of structured interfaces, weak articulation between knowledge types, and limited availability of shared data. Together, these findings indicate divergent expectations regarding what counts as relevant evidence and limit the stabilisation of shared evidentiary reference points. Knowledge is therefore not collectively interpreted or aligned, but remains segmented across actor-specific logics.

#### Pattern 6. Institutional preferences selectively filter what becomes actionable

Interviewees consistently highlight the political value of quantified data in contexts of scarce resources and competing health priorities. Numbers—particularly those related to costs, coverage, and population burden—are described as central to agenda-setting and arbitration.

Such data are often scarce, outdated, or inaccessible. In practice, quantification operates as a powerful filter: information that can be readily converted into cost estimates, coverage indicators, or comparable figures circulates more easily in decision arenas, while other forms of knowledge remain peripheral unless translated into measurable formats.

Experiential knowledge is widely recognised as powerful for mobilisation and public visibility, but is rarely sufficient, on its own, to support budgetary or strategic decisions. This limitation is not framed as a lack of legitimacy, but as a problem of translation: without articulation with quantified and synthesised data, lived experience remains weakly connected to the forms of evidence expected in arbitration arenas.

Consequently, evidence is not absent but selectively visible. What cannot be easily counted, compared, or summarised in concise formats is often sidelined from arbitration and planning processes. Institutional constraints—time pressure, workload, and hierarchical decision structures—further reinforce these dynamics by favouring concise, operational forms of evidence.

These preferences shape not only how evidence is presented, but also which knowledge is considered usable in practice. Interviewees converge on a relatively narrow set of elements perceived as actionable: key population figures, cost estimates, indicators of financial hardship, and simplified descriptions of care gaps. Experiential knowledge—although widely recognised as legitimate and valuable for mobilisation—remains weakly convertible into decision-oriented evidence in the absence of translation into these formats.

This filtering dynamics are reinforced by weakly structured processes of deliberation and prioritisation. in which evidence is not systematically synthesised, compared, or arbitrated. A recurring pattern emerges in which a small number of emblematic priorities—such as the creation of a national oncology centre or the coverage of treatments by the Assurance Maladie Obligatoire (AMO, mandatory health insurance scheme) and the Régime d’Assistance Médicale (RAMED, publicly funded medical assistance scheme for people identified as indigent and unable to pay for care) —are repeatedly invoked across policy windows. These priorities function as shared reference points, creating apparent alignment across actors, but rarely translate into sustained financing, routinised coordination, or clear implementation pathways. Rather than reflecting evidentiary convergence, this “surface consensus” corresponds to the discursive stabilisation of highly visible options within fragmented debates, narrowing deliberation around a limited set of symbolically powerful interventions.

This pattern corresponds to recurrent cross-construct combinations identified across interviews (C3-C to C3-E, S2 File), including institutional constraints, preferences for concise formats, limited translation of experiential knowledge, and weakly structured prioritisation processes. Combined, these elements show that the boundaries of actionable evidence are not fixed, but negotiated within decision-making arenas. Knowledge that aligns with dominant institutional expectations is more likely to be taken up, while other forms remain peripheral unless translated into institutionally legible formats. In this configuration, intermediation is constrained not only by the availability of evidence, but also by the criteria through which evidence is recognised as relevant and usable.

### Cross-cutting conditions shaping the possible development of an association-led knowledge transfer mechanism

Across the three configurations, the findings identify a set of interdependent conditions shaping the possible development of an association-led knowledge transfer mechanism for breast cancer in Mali. Conditions of emergence include minimal institutional interfaces between research, patient associations, and decision-making spaces, together with some degree of political and administrative recognition. Conditions of functioning relate not only to organisational capacities within patient associations, but also to access to data and analytical support, understanding of the health system and decision-making processes, and the ability to articulate experiential knowledge in forms that can be heard and used within institutional spaces, alongside complementary roles between associations, researchers, and public institutions. Conditions of continuity depend on the routinisation of interactions, the alignment of knowledge formats with decision-making expectations, and the progressive anchoring of intermediation functions beyond individual actors or external projects. Intermediation is unlikely to develop when only one dimension is addressed in isolation; rather, institutional, organisational, and epistemic conditions need to be aligned jointly.

## Discussion

This study contributes to the KT literature by examining the conditions under which patient associations may contribute to intermediation in breast cancer governance in Mali. While KT researches has documented the role of brokers and boundary-spanning arrangements, patient associations remain comparatively under-theorised in this function, particularly in LMIC cancer contexts. Our findings suggest that their intermediary potential cannot be understood through associative characteristics alone, but depends on how organisational capacities, institutional continuity, and the legitimacy of different forms of knowledge are articulated within the governance environment.

### From potential intermediary assets to conditional knowledge intermediation in cancer governance

Our point of departure was that, in a context where no actor routinely assumes this function, patient associations appeared to hold several characteristics that could make them relevant contributors to knowledge intermediation: proximity to lived experience, public credibility, mobilisation capacity, and the ability to make otherwise neglected care realities visible. The findings confirm the relevance of these assets, but also show that they do not in themselves produce a stabilised intermediary role. Whether such a role can emerge depends on how these assets are connected to organisational capacities, institutional interfaces, and accepted formats of evidence use.

Patient associations do not only mobilise attention or provide testimony. They also generate distinctive forms of situated knowledge by rendering visible lived care trajectories, barriers to diagnosis and treatment, discontinuities in care, quality gaps, implementation bottlenecks, the social and economic consequences of illness, and the frictions patients encounter when navigating the health system. Their contribution to KT therefore extends beyond awareness-raising to the identification, collectivisation, framing, and communication of experiential knowledge in ways that can inform both service delivery debates and policy discussions.

Existing literature helps situate these findings. In both HIC and LMIC settings, the contribution of affected groups to knowledge use and policy influence depends less on experiential proximity alone than on the arrangements through which this experience is translated, legitimised, and connected to decision-making processes. Evidence from the HIV field in LMICs is particularly instructive: community-led monitoring shows that affected communities can contribute to actionable knowledge and policy dialogue when experiential and community-generated data are supported by structured accountability mechanisms, programme interfaces, and sustained capacity strengthening (35–37). In HICs, earlier work on AIDS activism and breast cancer advocacy similarly showed that affected groups became influential through processes of credibility-building, alliance formation, training, and negotiated access to research and policy arenas (38–42).

Our findings point in the same direction, while also highlighting specific features of cancer governance. Intermediation emerges as a distributed function across actors, with patient associations playing a distinctive role by mobilising experiential knowledge, conferring social legitimacy on issues, connecting institutional processes with lived realities, and helping to repeatedly bring attention back to a condition that remains weakly prioritised in policy and resource allocation. In parallel, cancer governance remains strongly associated with specialised, technology-intensive, and hospital-centred models of care. Decision-making unfolds across multiple interconnected arenas—clinical, administrative, and budgetary—in which the boundaries of legitimate and actionable knowledge are negotiated and tend to favour quantitative and biomedical formats (43,44). In this context, intermediation involves not only making experiences visible or disseminating research findings, but articulating heterogeneous forms of knowledge and navigating across intertwined decision-making levels.

The practical implication is not that patient associations should simply be “included more” or expected to assume a fully fledged intermediary role on their own. Rather, the findings point to the need to make their contribution credible and operational.

While patient associations hold a certain degree of legitimacy in mobilisation and awareness-raising, they still face challenges in establishing credibility in the production and mobilisation of evidence derived from both experiential and research-based knowledge. This remains particularly difficult in a context where patient participation is only weakly institutionalised. This gap is consistent with the 2008 Patient Charter (34), which codifies fundamental rights within health facilities, including access to care, information, and respect for dignity, but does not establish a structured role for patients or patient associations in health decision-making. National health policy is similarly grounded in principles of equity, solidarity, and participation, yet these provisions remain weakly institutionalised and are rarely mobilised in practice (45). Even the Charter acknowledges that codifying rights is insufficient without concrete measures to ensure that they are known, implemented, and claimed. As a result, patient participation remains symbolic, with limited translation into structured roles or sustained involvement in decision-making processes.

Beyond issues of credibility, several conditions need to be met to enable the operationalisation of an intermediary role for patient associations. These include support for the collectivisation and formalisation of experiential knowledge; earlier and more continuous spaces for joint interpretation between associations, researchers, clinicians, and decision-makers; and coordination arrangements capable of translating heterogeneous forms of knowledge into institutionally usable inputs. The contribution of patient associations therefore depends less on their intrinsic characteristics than on the arrangements that enable their role to become credible, usable, and sustained in practice and on processes through which individual experiences can be collectivised, stabilised, and translated into actionable knowledge in policy spaces (41,36).

These findings call for a reconsideration of how patient participation is conceptualised in cancer knowledge ecosystems. In highly technocratic and biomedically oriented governance settings, expanding the role of patient associations towards knowledge intermediation also raises questions about the distribution of epistemic authority. It reflects broader hierarchies in the valuation of knowledge, in which forms of knowledge rooted in embodied experience and social relations—often associated with women—remain less easily stabilised, formalised, and recognised as actionable evidence within institutional processes. Experiential legitimacy thus operates as both a resource and a constraint: it enables mobilisation and visibility, while limiting the capacity of associations to translate lived experience into collectively stabilised and policy-relevant forms of knowledge.

This tension resonates with broader analyses of women’s public and associative participation in Mali, where women’s organisations often constitute important spaces of mobilisation, social legitimacy, and political emergence, while women’s influence in formal decision-making arenas remains constrained (46). Breast cancer patient associations may therefore face a form of double marginalisation within decision-making arenas: first as bearers of experiential knowledge that remains weakly institutionalised, and second as women-led collective actors whose contribution is more readily recognised in registers of mobilisation than in those of policy-oriented expertise.

Patient associations can therefore link different forms of knowledge and legitimacy, but only if supported by mechanisms that enable their articulation across institutional spaces. Without such arrangements, they risk remaining highly visible in public or advocacy arenas while remaining marginal in strategic and budgetary decision-making.

### Designing KT interventions around functions, interactions, and absorption

A central implication of this study concerns what these findings mean for the design of public health action in breast cancer governance. While patient associations align with several recognised priorities for effective KT—such as integrating experiential knowledge, mobilising trusted actors, and promoting interactive processes (19,21) —these assets are unlikely to produce sustained effects when KT remains organised around a sequential division of labour. In such arrangements, associations mobilise, researchers produce knowledge, and decision-makers intervene only at the stage of validation, limiting joint interpretation and weakening the translation of knowledge into operational orientations.

The challenge is therefore not to redefine mandates or treat patient associations as standalone intermediaries, but to design KT interventions around the functions that need to be performed and how they are distributed across actors. This implies creating arrangements through which experiential mediation, involvement in research agenda-setting and evidence generation, contextual translation, convening, and policy alignment can be jointly articulated (16,19,21,47). It also requires engaging actors earlier and more continuously in shared spaces of interpretation, rather than connecting them only at the final stage.

In this perspective, KT interventions may be more effective when conceived not only as mechanisms for circulating knowledge, but as arrangements that support its absorption. The main issue identified in this study was not insufficient transmission of evidence, but weak organisational and institutional conditions for interpreting and acting upon it. Efforts focused solely on training, tools, or platforms are therefore unlikely to be sufficient if they are not embedded in the routines, relationships, and governance mechanisms through which knowledge becomes usable in practice (43,44).

From a public health perspective, this points to several practical directions: creating regular spaces for joint interpretation between patient associations, researchers, clinicians, and decision-makers; involving patient associations earlier in research processes so that experiential concerns can help shape research questions and evidence generation; developing concise, decision-oriented knowledge products that combine quantified evidence with structured experiential insights; and supporting patient associations in the collectivisation and translation of lived experience into policy-relevant forms. In the Malian context, such efforts are likely to be more effective when embedded within existing governance structures rather than organised as parallel mechanisms.

These findings also have implications for donors and public institutions. Strengthening KT requires attention not only to individual and organisational capacities, but also to the organisation of interactions, the alignment of roles and expectations, and the integration of KT processes within existing governance arrangements. In fragmented and resource-constrained settings, interventions that do not engage with these dimensions are unlikely to achieve sustained effects (15,43,44).

### The prospective and configuration-based use of CFIR provides a formative baseline for designing and evaluating association-led intermediation

Beyond its empirical findings, this study contributes to methodological discussions on the use of the Consolidated Framework for Implementation Research (CFIR) in global public health. While CFIR is most often applied retrospectively to analyse implemented interventions, it was used here prospectively to examine the conditions under which a knowledge transfer mechanism involving patient associations could plausibly emerge, function, and consolidate within a constrained health system.

The analysis combined construct-based coding with a configuration-oriented approach. Rather than analysing determinants in isolation, recurrent clusters of co-occurring constructs were identified across interviews and interpreted in terms of how their interactions shaped knowledge use and intermediation. This made it possible to identify recurrent patterns arising from these interactions and to group them into broader configurations according to their effects on the emergence, functioning, and durability of the mechanism.

A construct-by-construct analysis would have highlighted familiar implementation barriers—limited resources, fragmented governance, weak evidence use routines, and capacity constraints within patient associations—but would have provided limited insight into how these determinants combine to shape implementation conditions. By contrast, the configurational analysis highlights three interrelated dynamics: the structural discontinuity of knowledge use within fragmented and project-dependent governance arrangements; the gap between the recognised legitimacy of patient associations and their limited capacity to assume intermediation functions; and the negotiated boundaries of actionable evidence within policy arenas.

The prospective use of CFIR further allows these dynamics to be interpreted as structuring conditions shaping the feasibility of intermediation, rather than as barriers identified ex post. In this perspective, they point to priority domains for adaptive design and monitoring, including institutional interfaces, organisational capacities, actors’ positioning in relation to knowledge, and the routinisation of interactions.

Our research suggest that CFIR can be extended beyond its conventional retrospective use to support the analysis of pre-implementation feasibility and institutional positioning. Combined with a configuration-oriented perspective, it provides a structured basis for identifying how interacting determinants shape implementation conditions and for guiding the design and evaluation of knowledge transfer mechanisms in institutionally constrained settings.

## Strengths and limitations

This study has several limitations. First, it is based on qualitative data collected within a single national context and focused on breast cancer governance, which may limit transferability to other policy domains or institutional environments. Although the patterns identified resonate with analyses from other low- and middle-income settings, their applicability should be interpreted cautiously. In particular, perspectives from rural areas and from women facing the most severe geographic and socioeconomic barriers to cancer care may be under-represented. Second, interviews were conducted among actors directly involved in cancer governance and research processes. As in all qualitative studies, accounts may reflect positional perspectives or strategic framing, despite efforts to triangulate across stakeholder groups. Future research will be needed to examine how proposed intermediary configurations evolve in practice and whether they lead to sustained changes in evidence use.

## Conclusion

By focusing on patient associations as potential intermediary actors, this study highlights that strengthening KT in cancer governance does not consist in adding an additional actor or solely reinforcing technical capacities. Rather, it requires acting on relational arrangements, institutional interfaces, and formats of deliberation that enable the articulation of scientific and experiential knowledge within prioritisation processes.

From this perspective, patient associations appear as relevant actors not only because of their proximity to affected populations, but also because of their potential to surface social issues that remain insufficiently addressed in arenas dominated by technical and budgetary considerations. However, this contribution remains conditional on favourable organisational, institutional, and political environments. More specifically, it depends on the stabilisation of institutional interfaces, the consolidation of organisational capacities within associations, and the existence of shared processes through which experiential and scientific knowledge can be jointly interpreted and recognised as actionable, while accounting for the gendered conditions that shape women’s engagement and the recognition of experiential knowledge.

## Ethical considerations

The SENOVIE Mali study protocol was submitted to, evaluated and approved by the ethics committee of the Institut National de Santé Publique (Bamako, Mali) on 26 October 2021 under No. 17/2021 /CE-INSP. All participants gave their written consent to take part in the study.

## Data availability

The qualitative data generated and analysed during this study are not publicly available due to confidentiality and the risk of participant re-identification. De-identified excerpts supporting the findings are included in the article and supplementary materials. Additional information may be available from the corresponding author upon reasonable request and subject to ethical approval.

## Acknowledgments

The authors thank all participants who contributed to this study through interviews, discussions, and participation in research and knowledge transfer activities. We are particularly grateful to the patient associations, healthcare professionals, researchers, and institutional actors who shared their experiences and perspectives throughout the research process.

The multi-country SENOVIE research group associates Clémence Schantz (scientific leader), Moufalilou Aboubakar, Myriam Baron, Gaëtan Des Guetz, Anne Gosselin, Pascale Hancart Petitet, Joseph Larmarange, Hamidou Niangaly, Beauta Rath, Luis Teixeira, Bakary Abou Traoré (co-scientific leader), Anthelme K. Agbodande, Mena Agbodjavou, Audrey Bochaton, Sarah Boisson, Emmanuel Bonnet, Tararath Bun, Fanny Chabrol, Abdourahmane Coulibaly, Karna Coulibaly, Justin Lewis Denakpo, Annabel Desgrées du Loû, Kadiatou Faye, Freddy Gnangnon, Sineath Hong, Vannarith Kao, Léa Prost Lançon, Kimsonphanuth Muy, Valéry Ridde, Julie Robin, Hélène Sacca, Laetitia Someil, Angéline Tonato Bagnan and Alassane Traoré.

Within SENOVIE, the Mali research group is composed of Hamidou Niangaly and Clémence Schantz (co-principal investigators of the cohort); Martine Audibert, Boureima Bélem, Abdourahmane Coulibaly, Karna Coulibaly, Idrissa Diarra, Fatou Diawara, Kadiatou Kanté, Aboubakary Konaté, Abdramane Alou Koné, Joseph Larmarange, Madani Ly, Charlotte Ngo, Moussa A. Ouattara, Julie Robin, Issiaka Sagara, Toumani Sidibé, Ibrahima Téguété, Luis Teixeira, Ibrahim Térera, Tiounkani Augustin Thera, Adégné Togo, Alassane Traoré, Bakary Abou Traoré, Cheick B. Traoré, Drissa Traoré and Solomane Traoré, as associated researchers; and Hawa Diakité, Digama Kassambara, M’Bamoussa Kayentao, Ibrahim Koné, Kadidiatou Koné, Zakari Saye, Minata Sylla, Soumaila Tangara and Djénéba Togola as field investigators.

## Supporting information

S1 File. Adaptation and operationalisation of the CFIR framework for a prospective, configuration-oriented analysis of an association-led knowledge transfer mechanism

S2 File. Supplementary File S2 - Mapping of CFIR constructs, empirical clusters, and configuration-level patterns

S3 File. Illustrative quotations by configuration and cross-construct pattern

S4 File. Researcher reflexivity and methodological rigor

